# The prevalence of SARS-CoV-2 infection and other public health outcomes during the BA.2/BA.2.12.1 surge, New York City, April-May 2022

**DOI:** 10.1101/2022.05.25.22275603

**Authors:** Saba A Qasmieh, McKaylee M Robertson, Chloe A Teasdale, Sarah G Kulkarni, Heidi Jones, David A. Larsen, John J. Dennehy, Margaret McNairy, Luisa N. Borrell, Denis Nash

**Affiliations:** Institute for Implementation Science in Population Health (ISPH), City University of New York (CUNY); New York, NY, USA; Department of Epidemiology and Biostatistics, Graduate School of Public Health and Health Policy, City University of New York (CUNY); New York, NY, USA; Department of Public Health, Falk College, Syracuse University, Syracuse, NY, USA; Department of Biology, Queens College, City University of New York, Queens, NY, USA; Center for GLobal Health and Division of General Internal Medicine, Weill Cornell Medicine, New York, NY, USA

## Abstract

**Background:** Routine case surveillance data for SARS-CoV-2 are incomplete, unrepresentative, missing key variables of interest, and may be increasingly unreliable for both timely surge detection and understanding the burden of infection and access to treatment.

**Methods:** We conducted a cross-sectional survey of a representative sample of 1,030 New York City (NYC) adult residents ≥18 years on May 7-8, 2022, when BA.2.12.1 comprised 47% of reported cases per genomic surveillance. We estimated the prevalence of SARS-CoV-2 infection during the preceding 14-day period (April 23-May 8), weighted to represent the 2020 NYC adult population. Respondents were asked about SARS-CoV-2 testing (including at-home rapid antigen tests), testing outcomes, COVID-like symptoms, and contact with SARS-CoV-2 cases. Based on responses, we classified individuals into three mutually exclusive categories of SARS-CoV-2 infection according to a hierarchical case definition as follows: confirmed (positive test with a provider), probable (positive at home rapid test), and possible (COVID-like symptoms and close contact with a confirmed/probable case). SARS-CoV-2 prevalence estimates were age- and sex-adjusted to the 2020 US population. Individuals with SARS-CoV-2 were asked about awareness/use of antiviral medications. We triangulated survey-based prevalence estimates with NYC’s official SARS-CoV-2 metrics on cases, hospitalizations, and deaths, as well as SARS-CoV-2 concentrations in wastewater for the same time period.

**Results:** An estimated 22.1% (95%CI 17.9%-26.2%) of respondents had SARS-CoV-2 infection during the two-week study period, corresponding to ∼1.5 million adults (95%CI 1.3-1.8 million). The official SARS-CoV-2 case count during the study period was 51,218. This 22.1% prevalence estimate included 11.4%, 6.5%, and 4.3% who met the confirmed, probable, and possible criteria of our case definition, respectively. Prevalence was estimated at 34.9% (95%CI 26.9%-42.8%) among individuals with co-morbidities, 14.9% (95% CI 11.0%-18.8%) among those 65+ years, and 18.9% (95%CI 10.2%-27.5%) among unvaccinated persons. Hybrid immunity (i.e., history of both vaccination and prior infection) was 66.2% (95%CI 55.7%-76.7%) among those with COVID and 46.3% (95%CI 40.2-52.2) among those without. Among individuals with COVID, 44.1% (95%CI 33.0%-55.1%) were aware of the antiviral nirmatrelvir/ritonavir (Paxlovid™), and 15.1% (95%CI 7.1%-23.1%) reported receiving it. Deaths and hospitalizations increased, but remained well below the levels of the BA.1 surge. SARS-CoV-2 virus concentrations in wastewater surveillance showed only a modest signal in comparison to that of the BA.1 surge.

**Conclusions and Relevance:** The true magnitude of NYC’s BA.2/BA.2.12.1 surge may have been vastly underestimated by routine SARS-CoV-2 case counts and wastewater surveillance. Hybrid immunity, bolstered by the recent BA.1 surge, likely limited the impact of the BA.2/BA.2.12.1 surge on severe outcomes. Representative surveys are needed as part of routine surveillance for timely surge detection, and to estimate the true burden of infection, hybrid immunity, and uptake of time-sensitive treatments among those most vulnerable to severe COVID.

**Short abstract:** Changes in testing practices and behaviors, including increasing at-home rapid testing and decreasing provider-based testing make it challenging to assess the true prevalence of SARS-CoV-2. We conducted a population-representative survey of adults in New York City to estimate the prevalence of SARS-CoV-2 infection during the BA.2./BA.2.12.1 surge in late April/early May 2022. We triangulated survey-based SARS-CoV-2 prevalence estimates with contemporaneous city-wide SARS-CoV-2 metrics on diagnosed cases, hospitalizations, deaths, and SARS-CoV-2 concentration in wastewater. Survey-based prevalence estimates were nearly 30 times higher than official case counts, and estimates of recently acquired hybrid immunity among those with active infection were high. We conclude that no single data source provides a complete or accurate assessment of the epidemiologic situation. Taken together, however, our results suggest that the magnitude of the BA.2/BA.2.12.1 surge was likely significantly underestimated, and high levels of hybrid immunity likely prevented a major surge in BA.2/BA.2.12.1-associated hospitalizations/deaths.

## INTRODUCTION

Major surges in SARS-CoV-2 transmission, due to new or evolving variants and waning population immunity, are expected in many parts of the world for the foreseeable future. Depending on variant properties and the timing of surges in relation to population immunity, the impact of surges could be severe even in highly vaccinated populations. The Omicron (BA.1) surge in the U.S., beginning mid-December 2021 when 62% of the US population was fully vaccinated, overwhelmed the health care system and resulted in more than 187,000 deaths during a 4-month period.^1,2^ In the current phase of the pandemic, key components of the U.S. strategy to limit the impact of SARS-CoV-2 surges are vaccinations, timely boosters and, for those most vulnerable, prophylaxis with monoclonal antibodies, and rapid treatment with oral antivirals or monoclonal antibodies, which can greatly reduce the risk of severe disease and death (i.e., secondary prevention).^3–5^ As the pandemic progresses, levels of hospitalization and death among those most vulnerable to severe COVID-19 are likely to vary by locality, and to be influenced by variant properties (transmissibility, severity, immune evasion), population levels of immune protection (via vaccination/boosters and/or prior infection), intervals between surges, and access to treatment (antivirals, monoclonal antibodies).

The complex and evolving nature of the US pandemic has led to calls for more robust, timely, and representative approaches to public health surveillance.^6^ Routine passive surveillance to inform the public health response relies on healthcare providers, testing providers, and laboratories to report data on those who are tested. While these surveillance data have been essential for tracking and responding to the COVID-19 pandemic, routinely reported testing data have become increasingly unreliable for timely surge detection and gauging the overall surge magnitude and sub-population burden.^6–8^ For example, during the latter half of NYC’s initial Omicron surge (BA.1), official case counts were likely 3-4 times lower than an estimate of infections from a representative sample of the adult population.^9^ Data from traditional (passive) surveillance underestimate the true burden of infection in the general population due to undiagnosed/untested cases,^10^ underreporting of cases by providers and labs, as well as the expanding use of at-home rapid antigen tests, which are not reflected in routine case surveillance in the U.S.^7,9,11^ Moreover, while SARS-CoV-2 provider and laboratory reporting is believed to be incomplete^6^, the extent of incomplete reporting has not been systematically evaluated in NYC or nationally, and may be influenced by surges in transmission, testing demand, or both. The degree of underdiagnosis and underreporting is likely differential by geographic and sociodemographic factors and variable over time^12,13^, and may prevent or delay surge detection, limiting the ability of individuals and governments to take precautions.

Surveillance data are also limited with regard to key information about cases such as race/ethnicity, vaccination status, history of prior SARS-CoV-2 infection, comorbidities and uptake of biomedical interventions such as oral antivirals and monoclonal antibodies. The lack of such information prevents systematic assessment of both the burden of infection and uptake of biomedical interventions among those who may be most vulnerable to a severe outcome.

Population-based surveys have been used as part of routine public health surveillance in the United Kingdom^14^ and NYC^15^ and can provide rapid and complementary information that addresses some of the limitations of traditional surveillance of SARS-CoV-2 cases, hospitalizations, and deaths.^6^

This study aimed to use an efficient and pragmatic survey-based approach to assess the burden of SARS-CoV-2 infection during the BA.2/BA.12.2.1 surge in NYC starting in March 2022. In addition to prevalence data, the survey captured data on clinical outcomes, and the intersection of vaccine-induced and infection-induced immunity. We triangulated information from our survey with official counts of cases, hospitalizations, and deaths from routine surveillance and with data on SARS-CoV-2 concentration in NYC wastewater for the same time period.

## METHODS

### Survey-based estimation of SARS-CoV-2 prevalence

We conducted a cross-sectional survey, in English and Spanish, during May 7-8, 2022, of 1,030 adult New York City (NYC) residents via landlines (IVR) and mobile phones (SMS text). Potential participants were randomly selected from a sampling frame. Additional details on the survey design and sampling are in Appendix 1. Respondents were asked about SARS-CoV-2 testing and related outcomes during the 14 days prior to the survey (April 23-May 8). During the same time period, the BA.2.12.1 subvariant rose from an estimated 32% of reported cases to 47%.^16^

The study protocol was approved by the Institutional Review Board at the City University of New York (CUNY).

#### Point Prevalence Estimation

The survey questionnaire (Appendix 2) ascertained SARS-CoV-2 testing, including the location, types and results of viral diagnostic tests taken in the 14 days prior to the survey (PCR, rapid antigen and/or at-home rapid tests). The survey also captured information on COVID-19 symptoms, as well as known close contacts with a confirmed or probable case of SARS-CoV-2 infection. COVID-19 symptoms included any of the following: fever of ≥100°F, cough, runny nose and/or nasal congestion, shortness of breath, sore throat, fatigue, muscle/body aches, headaches, loss of smell/taste, nausea, vomiting and/or diarrhea.^17^ Participants were also asked about vaccination status, comorbidities that increase vulnerability to severe COVID-19, and prior history of SARS-CoV-2 infection/COVID. Participants who reported any type of COVID-19 test with a healthcare or testing provider, regardless of the result, were asked about awareness and uptake of the antiviral nirmatrelvir/ritonavir oral tablets (Paxlovid™), which NY State made available in the Spring of 2022.

Information gathered from respondents was used to estimate the number and proportion of respondents who likely had SARS-CoV-2 infection during the study period based on the following mutually exclusive, hierarchical case classification: 1) Confirmed case: self-report of one or more positive tests with a health care or testing provider; or 2) Probable case: self-report of a positive test result exclusively on at-home rapid tests (i.e. those that were not followed up with confirmatory diagnostic testing with a provider); or 3) Possible case: self-report of COVID-like symptoms AND a known epidemiologic link (close contact) to one or more laboratory confirmed or probable (symptomatic) SARS-CoV-2 case(s)^17^ in a respondent who reported never testing or only testing negative during the study period.

#### The intersection of vaccine- and infection-induced immunity

We combined information on vaccination status with that on prior COVID infections. Those who were fully vaccinated and those who were also boosted (fully vaccinated/boosted) with a history of prior COVID were classified as having, ‘hybrid immunity’ against severe COVID-19; those who were fully vaccinated or boosted with no history of prior COVID were classified as having ‘vaccine-induced immunity only’; those who were not fully vaccinated but had a history or prior COVID were classified as having ‘infection-induced immunity only’; and those who were neither vaccinated/boosted nor had a history of COVID were classified as having ‘no prior immunity’ (SARS-CoV-2 näive).

#### Statistical Analysis

We estimated the prevalence of SARS-CoV-2 by socio-demographic characteristics, NYC borough (county), vaccination status, comorbidity and prior COVID-19 infection status. Survey weights were applied to generate weighted numbers and estimates of the proportion who had active SARS-CoV-2 infection at any time during the study period along with 95% confidence interval (95%CI). We applied these weighted sample proportions and 95% CIs to the 6,740,580 NYC residents ≥18 years to obtain estimates of the absolute number of adults with SARS-CoV-2 infection.^18^ We used direct standardization to present age and sex adjusted prevalence estimates using the US 2020 census. Crude and age and sex-adjusted prevalence ratios were estimated with a log-binomial model. Pearson’s chi-squared test was performed to assess associations between each factor and testing status. Analyses were conducted using SAS and R.

### SARS-CoV-2 routine testing and case surveillance data

We used publicly available, daily aggregated data on the number of SARS-CoV-2 tests, test types (PCR or rapid antigen), and results through June 10, 2022 to describe the number of tests and positive tests with health care providers and testing providers reported to the NYC DOHMH during the study period.^19^

### SARS-CoV-2 wastwater surveillance data

We analyzed publicly available data on SARS-CoV-2 concentrations in NYC wastewater through June 5, 2022, which is estimated based on weekly influent samples from 14 water resource recovery facilities (WRRFs) in NYC covering wastewater of an estimated 8.2 million residents.^20^ Specifically, available data included the weekly per capita SARS-CoV-2 load (N1 copies per day per population), which represents “the total number of SARS-CoV-2 viruses per capita in wastewater entering a WRRF over a 24 hour period”.^20^ We plotted the mean per capita SARS-CoV-2 load (N1 copies per day per population) for all 14 WRRFs. Details on sampling and laboratory methods and measurement are available in the public use dataset documentation.^20^

## RESULTS

### Survey

An estimated 22.1% (95%CI 17.9%-26.2%) of respondents had SARS-CoV-2 infection in the 14 days prior to the interview, corresponding to 1.5 million adults (95% CI 1.3-1.8 million) (Table 1). The estimate of 22.1% includes: 1) 11.4% (95%CI 8.4%-14.3%) who were positive based on one or more tests with a health care or testing provider (confirmed cases); 2) 6.5% (95%CI 4.2%-8.8%) who were positive exclusively based on one or more positive at-home rapid tests (probable cases); and 3) 4.2% (95%CI 1.8%-6.7%) who met the definition for possible SARS-CoV-2 infection based on having *both* COVID-like symptoms and a close contact with a confirmed/probable case. About 53.8% of adults in our survey reported having any SARS-CoV-2 test during the study period, including 43% who reported testing with a health care or testing provider (5.5% exclusively) and 48% who tested using an at-home rapid test (10.9% exclusively).

**Table 1.** Characteristics for survey respondents by testing status and point prevalence of SARS-CoV-2, NYC April-May 2022

|  | Total | Crude prevalence of SARS-CoV-2 infection† | Age and sex direct-standardized prevalence of SARS-CoV-2 infection** | Crude prevalence ratio (PR) | Adjusted prevalence ratio (aPR)*** |
| --- | --- | --- | --- | --- | --- |
|  | Weighted N (%) | % (95% CI) | % (95% CI) (Weighted N = 991) | PR (95% CI) (Weighted N = 1,030) | aPR (95% CI) (Weighted N = 1,030) |
| **Total** | 1030 (100) | 22.1 (17.9 - 26.2) |  |  |  |
| **Testing** |  |  |  |  |  |
| Confirmed | 117 (11.4) | 11.4 (8.4 - 14.3) |  |  |  |
| Probable | 67 (6.5) | 6.5 (4.2 - 8.8) |  |  |  |
| Possible cases (non-testers/negatives) | 44 (4.3) | 4.2 (1.8 - 6.7) |  |  |  |
| **Age** |  |  |  |  |  |
| 18-24 | 112 (10.8) | 27.7 (12.7-42.8) | 26.1 (14.2 - 42.9) | 1.3 (0.9 - 1.9) | 1.4 (0.9 - 2.00) |
| 25-34 | 232 (22.5) | 21.6 (12.0 - 31.1) | 21.4 (13.4 - 32.4) | -ref- | -ref- |
| 35-44 | 176 (17.1) | 20.5 (8.1 - 32.8) | 21.1 (11.0 - 36.6) | 0.9 (0.6 - 1.4) | 1.0 (0.7 - 1.4) |
| 45-54 | 159 (15.5) | 27.8 (15.9 - 39.7) | 28.0 (17.7 - 41.2) | 1.3 (0.9 - 1.8) | 1.3 (0.9 - 1.9) |
| 55-64 | 158 (15.3) | 23.6 (15.4 - 31.9) | 22.4 (15.3 - 31.6) | 1.1 (0.8 - 1.6) | 1.1 (0.8 - 1.6) |
| 65+ | 194 (18.8) | 14.9 (11.0 - 18.8) | 13.7 (10.4 - 17.9) | 0.7 (0.5 - 1.0) | 0.7 (0.5 - 1.1) |
| **Gender** |  |  |  |  |  |
| Male | 471 (45.7) | 23.4 (16.5 - 30.2) | 22.8 (17.0 - 30.0) | 1.2 (0.9 - 1.5) | 1.1 (0.9 - 1.5) |
| Female | 527 (51.2) | 20.1 (14.9 - 25.3) | 20.1 (15.4 - 25.8) | -ref- | -ref- |
| Non-binary**** | 32 (3.1) | 36.1 (13.2 - 59.1) | 29.1 (15.2 - 48.6) | 1.8 (1.1 - 2.9) | 1.9 (1.1 - 3.1) |
| **Race/Ethnicity** |  |  |  |  |  |
| Black NH | 214 (20.8) | 11.3 (4.9 - 17.6) | 11.4 (6.7 - 18.7) | 0.4 (0.3 - 0.6) | 0.4 (0.3 - 0.6) |
| White NH | 393 (38.2) | 26.3 (20.0 - 32.5) | 26.0 (20.5 - 32.4) | -ref- | -ref- |
| Hispanic | 258 (25.0) | 33.6 (23.4 - 43.9) | 31.1 (22.6 - 41.1) | 1.3 (1.0 - 1.6) | 1.3 (1.0 - 1.7) |
| Asian/Pacific Islander | 121 (11.7) | 4.9 (0.0 - 10.5) | 5.2 (1.7 - 14.8) | 0.2 (0.1 - 0.4) | 0.2 (0.1 - 0.4) |
| Other | 45 (4.3) | 17.1 (0.0 - 35.9) | 14.1 (5.3 - 32.6) | 0.7 (0.3 - 1.3) | 0.6 (0.3 - 1.2) |
| **Years of education** |  |  |  |  |  |
| Some HS and below | 164 (16.0) | 28.8 (15.8 - 41.8) | 31.3 (20.8 - 44.2) | 1.3 (0.9 - 1.9) | 1.4 (1.0 - 2.1) |
| HS Grad | 239 (23.2) | 21.5 (12.3 - 30.8) | 20.0 (12.2 - 30.9) | -ref- | -ref- |
| Some college | 205 (19.9) | 16.7 (9.6 - 23.7) | 16.3 (10.6 - 24.5) | 0.8 (0.5 - 1.1) | 0.8 (0.5 - 1.2) |
| College grad and above | 422 (40.9) | 22.4 (16.3 - 28.5) | 23.1 (17.8 - 29.5) | 1.0 (0.8 - 1.4) | 1.0 (0.8 - 1.4) |
| **Household size** |  |  |  |  |  |
| 1 | 349 (33.8) | 16.7 (10.4 - 23.0) | 15.0 (9.8 - 22.6) | -ref- | -ref- |
| 2-3 | 403 (39.1) | 20.7 (14.8 - 26.7) | 21.1 (15.9 - 27.5) | 1.2 (0.9 - 1.7) | 1.2 (0.8 - 1.6) |
| 4+ | 278 (27.1) | 30.8 (20.9 - 40.6) | 25.7 (17.9 - 35.3) | 1.8 (1.4 - 2.5) | 1.7 (1.3 - 2.4) |
| **Any children <18 years** |  |  |  |  |  |
| Y | 271 (26.3) | 33.3 (23.0 - 43.5) | 31.5 (12.9 - 43.0) | 1.8 (1.5 - 2.3) | 1.7 (1.5 - 2.4) |
| N | 759 (73.7) | 18.1 (13.8 - 22.3) | 17.8 (13.6 - 22.8) | -ref- | -ref- |
| **Household income** |  |  |  |  |  |
| Below 25K | 221 (21.5) | 18.6 (10.0 - 27.2) | 14.2 (8.2 - 23.5) | 0.6 (0.4 - 0.8) | 0.6 (0.4 - 0.8) |
| 25,000 - 65,000 | 287 (27.9) | 24.9 (16.7 - 33.0) | 27.5 (19.3 - 37.5) | 0.8 (0.6 - 1.0) | 0.7 (0.6 - 1.0) |
| 65,000 - 150,000 | 229 (22.2) | 31.4 (20.8 - 42.1) | 32.3 (24.1 - 41.7) | -ref- | -ref- |
| Above 150,000 | 77 (7.5) | 17.9 (7.2 - 28.6) | 18.6 (10.5 - 30.8) | 0.6 (0.3 - 1.0) | 0.5 (0.3 - 0.9) |
| Prefer not to answer | 215 (20.9) | 13.5 (6.7 - 20.2) | 14.2 (7.8 - 24.4) | 0.4 (0.3 - 0.6) | 0.4 (0.3 - 0.6) |
| **Employed** |  |  |  |  |  |
| Yes | 477 (46.3) | 29.3 (22.2 - 36.5) | 30.9 (24.8 - 37.9) | -ref- | -ref- |
| No/DK | 553 (53.7) | 15.8 (11.4 - 20.3) | 13.4 (8.8 - 19.9) | 0.5 (0.4 - 0.7) | 0.5 (0.4 - 0.7) |
| **Nativity** |  |  |  |  |  |
| Born in U.S. | 714 (69.4) | 26.8 (21.3 - 32.2) | 26.5 (21.6 - 32.2) | -ref- | -ref- |
| Born outside U.S. | 316 (30.6) | 11.4 (6.6 - 16.2) | 10.8 (6.5 - 17.3) | 0.4 (0.3 - 0.6) | 0.4 (0.3 - 0.6) |
| **Borough** |  |  |  |  |  |
| Bronx | 175 (17.0) | 21.1 (10.4 - 31.8) | 20.2 (12.0 - 32.0) | 0.9 (0.6 - 1.4) | 0.9 (0.6 - 1.4) |
| Brooklyn | 317 (30.7) | 27.2 (19.8 - 34.7) | 26.1 (19.7 - 33.8) | 1.2 (0.9 - 1.6) | 1.2 (0.9 - 1.7) |
| Manhattan | 201 (19.5) | 22.6 (13.2 - 32.1) | 24.0 (15.5 - 35.1) | -ref- | -ref- |
| Queens | 278 (27.0) | 18.2 (10.4 - 26.1) | 17.6 (11.3 - 26.3) | 0.8 (0.6 - 1.2) | 0.8 (0.6 - 1.2) |
| Staten Island | 59 (5.7) | 13.6 (0.0 - 28.0) | 8.1 (2.8 - 20.9) | 0.6 (0.3 - 1.2) | 0.6 (0.3 - 1.3) |
| **Vaccination status** |  |  |  |  |  |
| Boosted | 691 (67.1) | 25.2 (19.8 - 30.5) | 25.2 (20.0 - 31.3) | 2.1 (1.4 - 3.4) | 2.1 (1.3 - 3.2) |
| Fully vaccinated not boosted | 149 (14.5) | 11.8 (3.5 - 20.1) | 11.8 (5.7 - 23.0) | -ref- | -ref- |
| Not vaccinated | 190 (18.5) | 18.9 (10.2 - 27.5) | 15.3 (9.6 - 23.5) | 1.6 (0.9 - 2.7) | 1.5 (0.9 - 2.5) |
| **Prior COVID since March 2020** |  |  |  |  |  |
| Never | 390 (37.9) | 10.7 (4.7 - 16.6) | 11.5 (6.7 - 18.9) | 0.3 (0.2 - 0.4) | 0.4 (0.4 - 0.6) |
| Once | 235 (22.8) | 39.2 (28.2 - 50.2) | 36.9 (27.6 - 47.3) | -ref- | -ref- |
| More than once | 140 (13.6) | 40.4 (27.2 - 53.6) | 37.5 (27.2 - 49.1) | 1.0 (0.8 - 1.3) | 1.0 (0.8 - 1.3) |
| Never tested positive but think they had COVID | 265 (25.8) | 14.1 (8.5 - 19.7) | 13.0 (8.2 - 20.0) | 0.4 (0.3 - 0.5) | 0.5 (0.4 - 0.7) |
| **Protection against severe disease** |  |  |  |  |  |
| Hybrid protection | 522 (50.6) | 28.9 (22.6 - 35.1) | 29.2 (23.1 - 36.0) | -ref- | -ref- |
| Vaccine-induced protection only | 318 (30.9) | 12.9 (5.6 - 20.1) | 12.9 (7.3 - 21.3) | 0.4 (0.3 - 0.6) | 0.4 (0.3 - 0.6) |
| Infection-induced protection only | 118 (11.5) | 29.8 (16.6 - 43.1) | 24.6 (16.4 - 35.2) | 1.0 (0.8 - 1.4) | 0.9 (0.7 - 1.3) |
| No protection | 72 (7.0) | 0.9 (0.0 - 2.7) | 1.7 (0.3 - 10.7) | 0.0 (0.0 - 0.4) | 0.0 (0.0 - 0.3) |
| **Comorbidities** |  |  |  |  |  |
| Y | 377 (36.6) | 34.9 (26.9 - 42.8) | 36.6 (28.3 - 45.8) | 2.4 (1.9 - 3.0) | 2.8 (2.2 - 3.5) |
| N | 653 (63.4) | 14.7 (10.4 - 19.0) | 14.7 (11.1 - 19.1) | -ref- | -ref- |
| **Any vulnerability\*** |  |  |  |  |  |
| Y | 609 (59.1) | 27.8 (22.1 - 33.5) | 29.6 (23.7 - 36.3) | 2.0 (1.5 - 2.7) | 2.5 (1.9 - 3.3) |
| N | 421 (40.9) | 13.8 (8.3 - 19.2) | 16.7 (11.4 - 23.7) | -ref- | -ref- |
| **Health insurance** |  |  |  |  |  |
| Yes | 836 (81.2) | 22.9 (18.2 - 27.6) | 22.6 (18.2 - 27.8) | -ref- | -ref- |
| No | 194 (18.9) | 18.5 (9.9 - 27.0) | 17.3 (10.6 - 27.0) | 0.8 (0.6 - 1.1) | 0.7 (0.5 - 1.0) |

\*Aged 65 or older OR $\geq 1$ comorbidity OR unvaccinated
\*\*Direct standardized for the age and sex groupings in the 2020 US census
\*\*\*Model adjusted for sex and age
\*\*\*\*Estimates are adjusted for age only

The weighted characteristics of survey participants and period prevalence estimates (both crude and age and sex-adjusted) are also shown in Table 1. In general, crude prevalence estimates and prevalence ratios were not materially altered by age- and sex-adjustment. SARS-CoV-2 prevalence was high among all groups, but varied substantially by sociodemographic factors, and was especially high among adults aged 18-24 (26.1%, 95%CI 14.2%-42.9%) and 45-54 (28.0%, 95%CI 17.7%-41.2%). Age and sex-adjusted prevalence was higher among Hispanic (31.1%, 95%CI 22.6% - 41.1%), and non-Hispanic White residents (26.0%, 95%CI 20.5%-32.4%), and those with some high school education or less (31.3%, 95%CI 20.8%-44.2%). Age and sex-adjusted prevalence estimates were the lowest among non-Hispanic Black (11.4%, 95%CI 6.7%-18.7%) and Asian/Pacific Islander (5.2%, 95%CI 1.7%-14.8%) residents. Age and sex-adjusted SARS-CoV-2 prevalence increased in dose response fashion with the number of household members, and households with children <18 years had substantially higher prevalence than those without (31.5% 95%CI 12.9%-43.0% vs 17.8%, 95%CI 13.6%-22.8%) (Figure 1).

**Figure 1.**
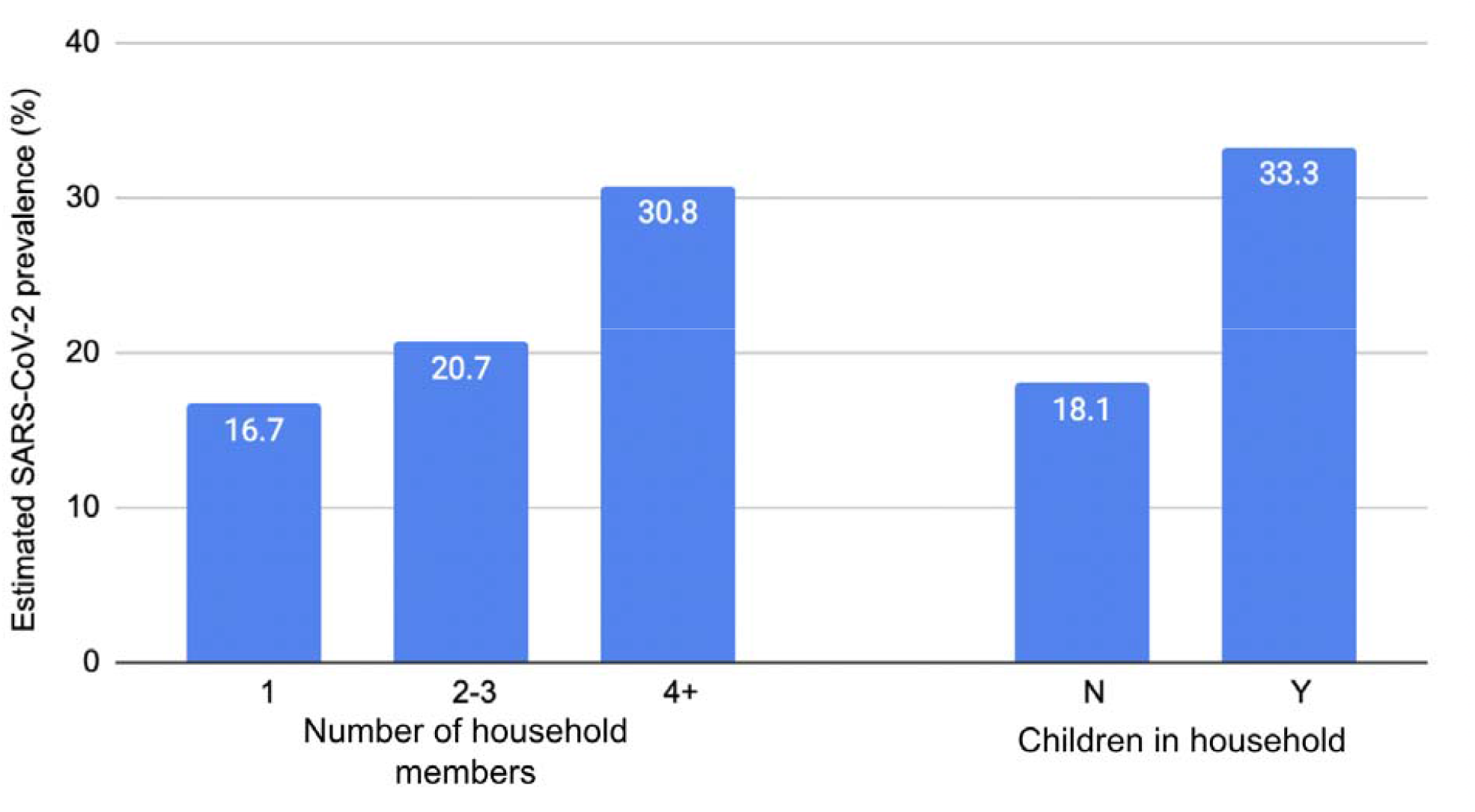
Estimated SARS-CoV-2 prevalence NYC adults by household size and presence of children in the household, April-May, 2022

Individuals who were fully vaccinated with a booster had higher age and sex-adjusted SARS-CoV-2 prevalence (25.2%, 95%CI 20.0%-31.3%) than those who were fully vaccinated but not boosted (11.8%, 95%CI 5.7%-23.0%) and those who were unvaccinated (15.3%, 95%CI 9.6%-23.5%). Those who said they tested positive for SARS-CoV-2 once (36.9%, 95%CI 27.6%-47.3%) or more than once (37.5%, 95%CI 27.2%-49.1%) had much higher age and sex-adjusted prevalence than those who said they never tested positive before (11.5%, 95% CI 6.7%-18.9%) or who thought they had COVID before but never tested positive (13.0%, 95%CI 8.2%-20.0%).

#### Hybrid immunity

Among those who were either vaccinated/boosted, those who also had SARS-CoV-2 infection in the past (hybrid immunity) had an age and sex-adjusted prevalence of 29.2% (95%CI 23.1%-36.0%), compared with 12.9% (95%CI 7.3%-21.3%) among those who did not have SARS-CoV-2 in the past (vaccine-induced protection only). Among those who were not vaccinated/boosted, those who had SARS-CoV-2 infection in the past (infection-induced immunity only) had an age-adjusted prevalence of 24.6% (95%CI 16.4%-35.2%), compared with 1.7% (95%CI 0.3%-10.7%) among those who did not have SARS-CoV-2 in the past (no prior SARS-CoV-2 immunity). The proportion of adults with hybrid immunity and infection-induced immunity only were higher in those with SARS-CoV-2 infection versus those without (Figure 2).

**Figure 2.**
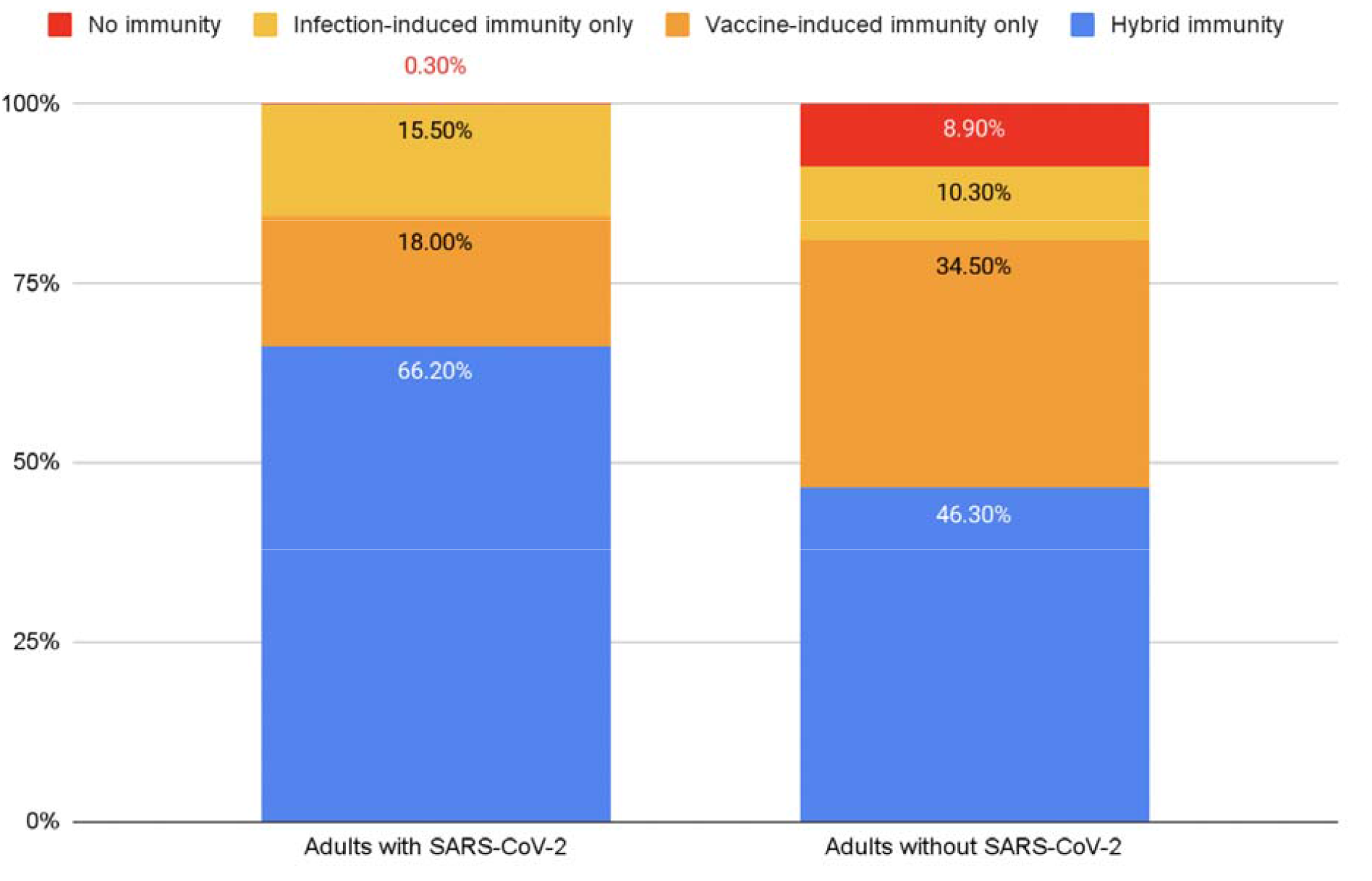
Hybrid immunity among adults with and without SARS-CoV-2 infection, NYC April-May, 2022

#### Vulnerability to severe COVID-19

The estimated prevalence of SARS-CoV-2 was substantial among groups vulnerable to severe SARS-CoV-2 and death, including unvaccinated persons (18.9%, 95%CI 10.2%-27.5%), those aged 65+ (14.9%, 95%CI 11.0%-18.8%), and individuals with co-morbidities (34.9%, 95%CI 26.9%-42.8%). Among those with any of these vulnerabilities to severe COVID-19 (age≥65, comorbidities, unvaccinated), 27.8% (95%CI 22.1%-33.5%) had SARS-CoV-2 infection. Also among this vulnerable group, only 68.8% (95%CI 62.8-74.8) were fully vaccinated with or without a booster. Specifically, 60.0% (95%CI 59.9-66.2) were fully vaccinated and boosted (of whom 68.8% [95%CI 61.5-76.2] had a history of prior COVID) and 8.7% (95%CI 5.6-11.8) were fully vaccinated but not boosted (of whom 71.8% [95%CI 54.4-89.1] had a history of prior COVID). However, 31.2% (95%CI 25.2-37.2) were unvaccinated (of whom 62.1% [95%CI 50.1-74.1] had a history of prior COVID).

#### Testing

Just over half of NYC adults (53.8%) reported any testing over the study period, including at-home testing (Table 2). Most of those testing reported testing with a provider (42.9%), corresponding to an estimated 2.9 million provider tests. Additionally, 10.9% of adults said they tested only at home. The characteristics of those who tested at all differed from those who did not test at all. Compared with those who did not test, those who tested were more likely to be younger, Hispanic, and non-Hispanic White, and to have: higher education, larger households, children in the household, lower household income, received a booster, had COVID more than once, hybrid and infection-induced immunity, medical vulnerabilities to severe COVID-19, and no insurance (Table 2).

**Table 2.**
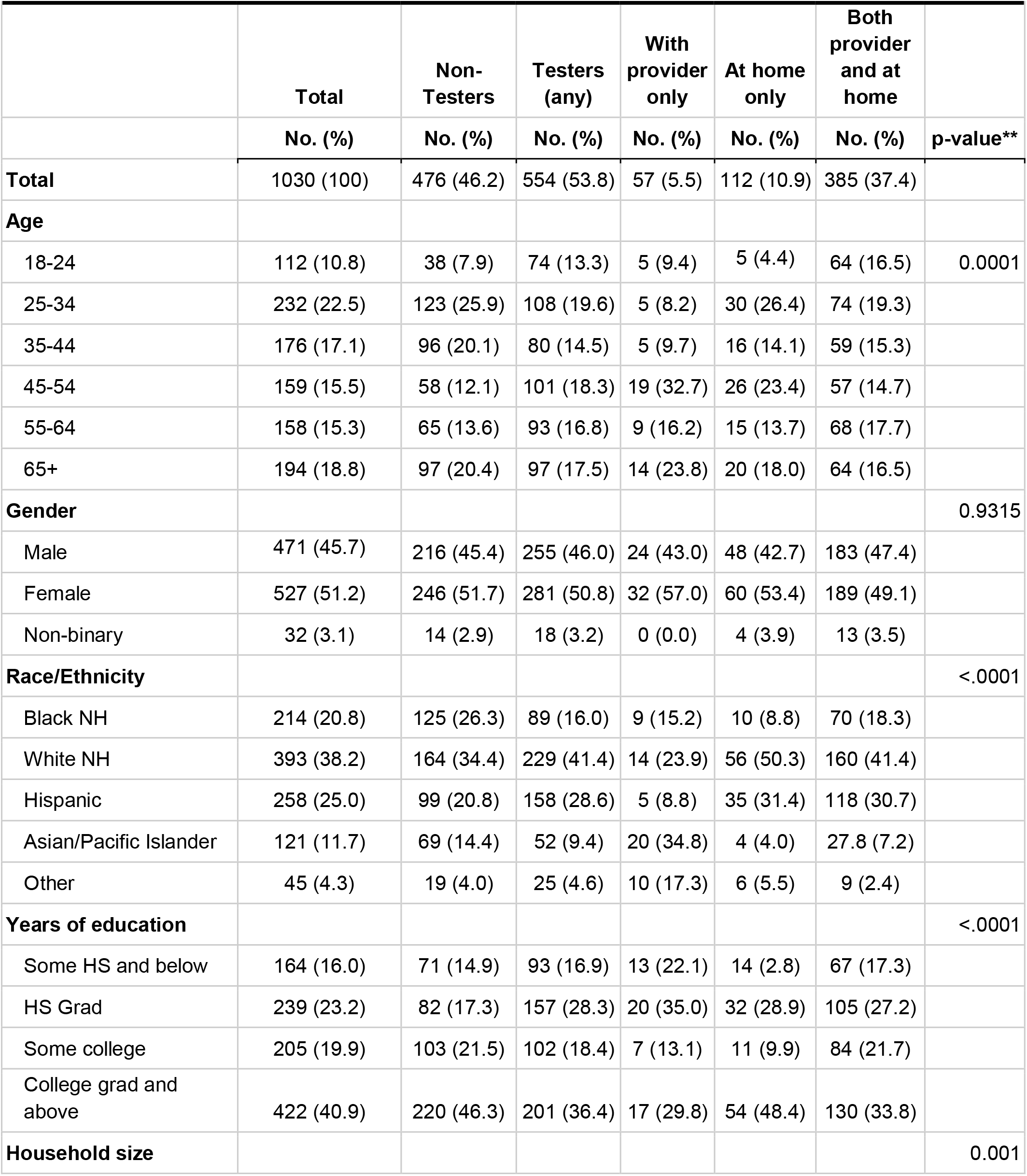
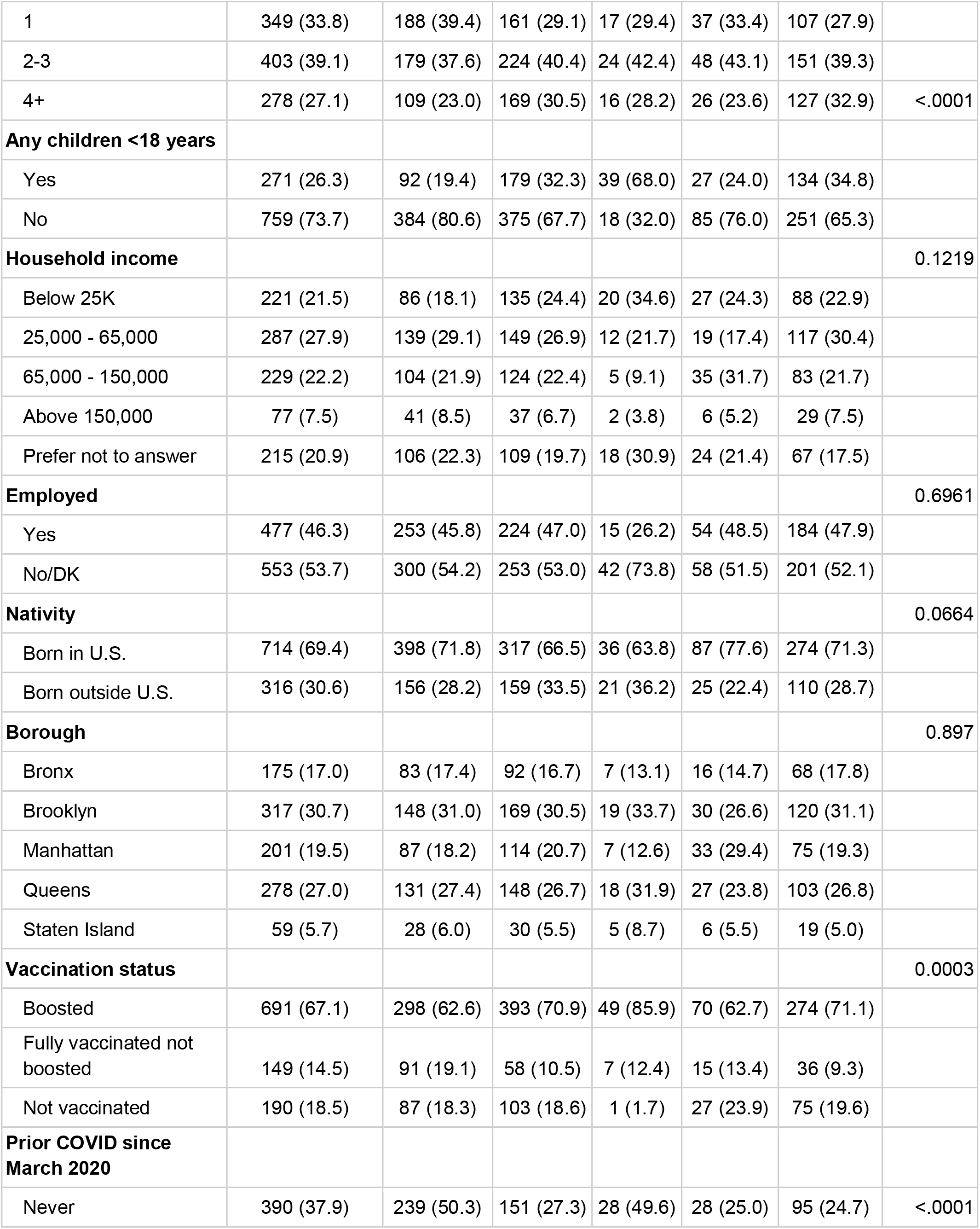
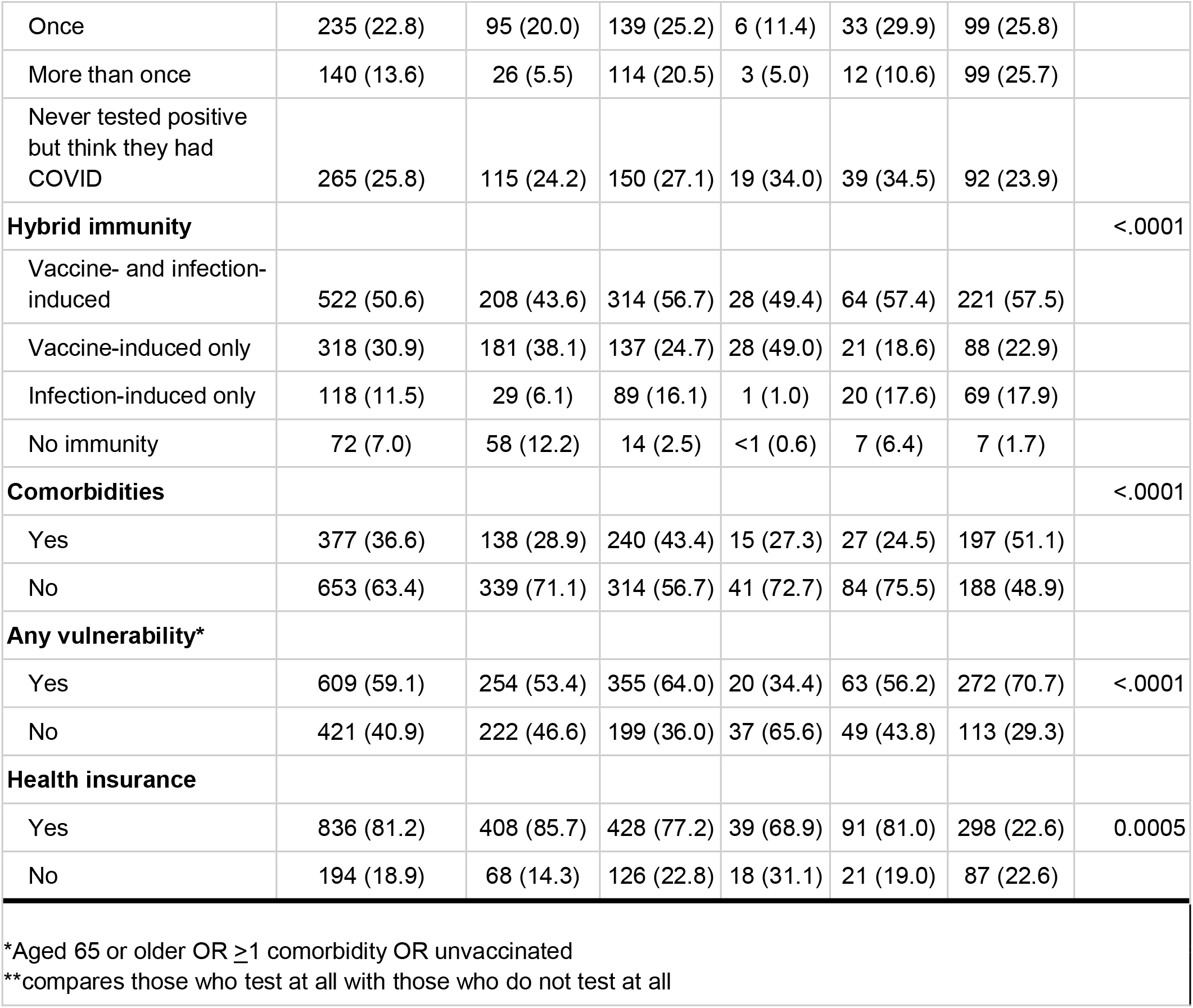
Characteristics of survey respondents by testing status, NYC April-May 2022

#### Vulnerability to severe COVID, awareness and uptake of antivirals among those with SARS-CoV-2 infection

Among the 22.1% with SARS-CoV-2 infection during the study period, 74.5% (95%CI 65.4-83.6) had one or more vulnerability, 66.1% (95%CI 55.7-76.7] had hybrid protection, and 29% (95%CI 19.6-38.6) met eligibility criteria for antivirals (by virtue of being aged 65+ or having one or more comorbidities)^21^ (Table 3). Of those who tested with a healthcare or testing provider, 55.9% (95%CI 44.9%-67.0%) were not aware of the antiviral nirmatrelvir/ritonavir and 3.0% (95%CI 0.0%-7.1%) reported that they tried to access it, but couldn’t.

**Table 3.** Characteristics and antiviral awareness and uptake among NYC adults with SARS-CoV-2 infection, April-May 2022

|  | **Total** |
| --- | --- |
|  | **No. (%)** |
| **Total number of positives** | 227 (100) |
| **Hybrid immunity** |  |
| Vaccine- and infection-induced | 151 (66.2) |
| Vaccine-induced only | 41 (18.0) |
| Infection-induced only | 35 (15.5) |
| No immunity | 1 (0.3) |
| **Vulnerability to severe COVID-19** |  |
| Any vulnerability* | 169 (74.5) |
| Unvaccinated | 36 (15.8) |
| Prior history of COVID | 35 (98.2) |
| No prior history of COVID | 1 (1.8) |
| Comorbidity* | 131 (57.8) |
| Age 65+ | 29 (12.7) |
| **Antiviral eligibility and awareness** |  |
| Eligible for antivirals** | 66 (29.0) |
| Awareness (n=192) |  |
| Unaware of antivirals | 108 (55.9) |
| Aware of antivirals | 84 (43.8) |
| Tried to access but couldn't | 6 (3.1) |

\*Aged 65 or older OR >1 comorbidity OR unvaccinated; possible comorbidities included: Cancer, diabetes, obesity, COPD or lung disease, liver disease, heart disease, high blood pressure, a recent organ transplant, or an immunodeficiency
\*\*Eligible: above 65 or with comorbidities, with reported symptoms and tested positive on at-home rapid or POC rapid or PCR test

Among those with a recent SARS-CoV-2 infection, 15.1% (95%CI 7.1%-23.1%) reported receiving nirmatrelvir/ritonavir, with substantial variability by sociodemographic groups (Table 4). Reported nirmatrelvir/ritonavir use was higher among those with any medical vulnerability versus those without, among non-Hispanic Whites and Asians compared with Hispanics and non-Hispanic Blacks, those with health insurance versus those without, those under 65 versus those older, those who were employed versus not, those who were college graduates versus those who were not, those with household income >25K versus those with lower income.

**Table 3.** Nirmatrelvir/ritonavir uptake*** among NYC adults with SARS-CoV-2 infection, April-May 2022

|  | **Total** |
| --- | --- |
|  | **% (95% CI)\*\*\*\*** |
| **Overall** | 15.1 (7.1- 23.1) |
| **Uptake by subgroup** |  |
| **Vulnerable to severe COVID-19** |  |
| Yes | 17.4 (7.5-27.3) |
| No | 6.9 (0.0-15.3) |
| **Age** |  |
| 65+ | 2.1 (0.0-5.4) |
| <65 | 17.2 (7.8-26.5) |
| **Age** |  |
| 55+ | 7.0 (0.13-13.9) |
| <55 | 18.7 (7.5-29.8) |
| **Race/ethnicity** |  |
| NH White | 19.7 (7.9-31.4) |
| NH Black | 6.5 (0.0-20.0) |
| Hispanic | 11.5 (0.0-25.7) |
| Asian | 52.7 (0.0-100) |
| **Health insurance** |  |
| Yes | 17.3 (7.9-26.7) |
| No | 3.8 (0.0-11.7) |
| **Employed** |  |
| Yes | 22.6 (10.1-35.2) |
| No | 2.9 (0.0-7.2) |
| **Education** |  |
| College graduate | 32.7 (16.5-48.8) |
| Not college graduate | 3.3 (0.0-7.1) |
| **Household income** |  |
| >25K | 17.8 (8.2-27.4) |
| ≤25K | 3.3 (0.0-10.2) |
| ***Uptake only assessed among 192 positive respondents who were in contact with a with a healthcare or testing provider |  |
| +Precision of estimates is low due to smaller sample size |  |

### Routine surveillance of SARS-CoV-2 testing, cases, hospitalizations and deaths

#### Testing and cases

Figure 3a. shows trends in SARS-CoV-2 tests administered by healthcare and testing providers with results reported to the NYC DOHMH among testers of all ages since the beginning of the pandemic through June 10, 2022. Throughout this entire period, 31.3 million PCR tests and 9.7 million rapid antigen tests were reported by providers and laboratories. PCR tests were the predominant test type reported (76%), comprising 83% of all positive tests overall. PCR tests remained predominant for the period since October 2020 when rapid antigen tests were more widely available at testing locations and comprised 73% of all tests and 80% of all positive tests. There were substantial declines in SARS-CoV-2 testing per 100,000 residents leading up to the study period (Figure 3a). During the study period (April 23-May 8, 2022), there were 671,377 tests reported, which gave rise to 51,218 cases (7.6% of tests). Of all tests, 505,242 (75%) were PCR tests, of which 33,066 (6.5%) were positive, and 166,135 (25%) were rapid antigen tests, of which 9,076 (5.5%) were positive.

**Figure 3a.**
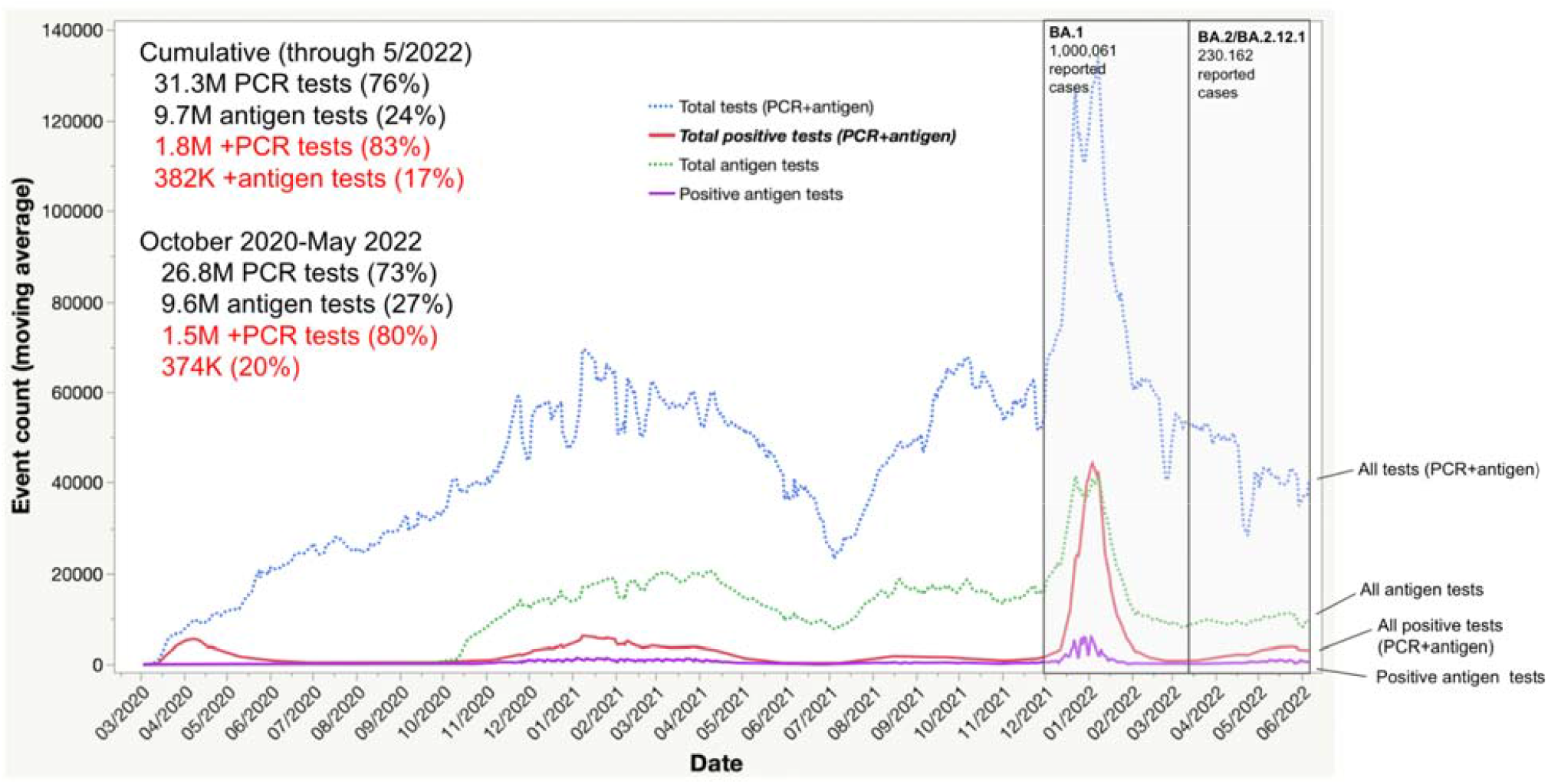
Total PCR tests, antigen tests, positive PCR tests, and positive antigen tests, NYC

#### Hospitalizations and deaths

COVID-19 hospitalizations and deaths increased, but modestly in comparison to that during the BA.1 surge during December 2021-February 2022 (Figure 3b). During December 1-March 14th, which brackets NYC’s BA.1 surge, there were 33,683 hospitalizations and 4,867 deaths. But during March 15th-June 20th, which includes the BA.2/BA.2.12.1 surge to date, the cumulative number of hospitalizations and deaths were much lower (6,983 hospitalizations and 651 deaths).

**Figure 3b.**
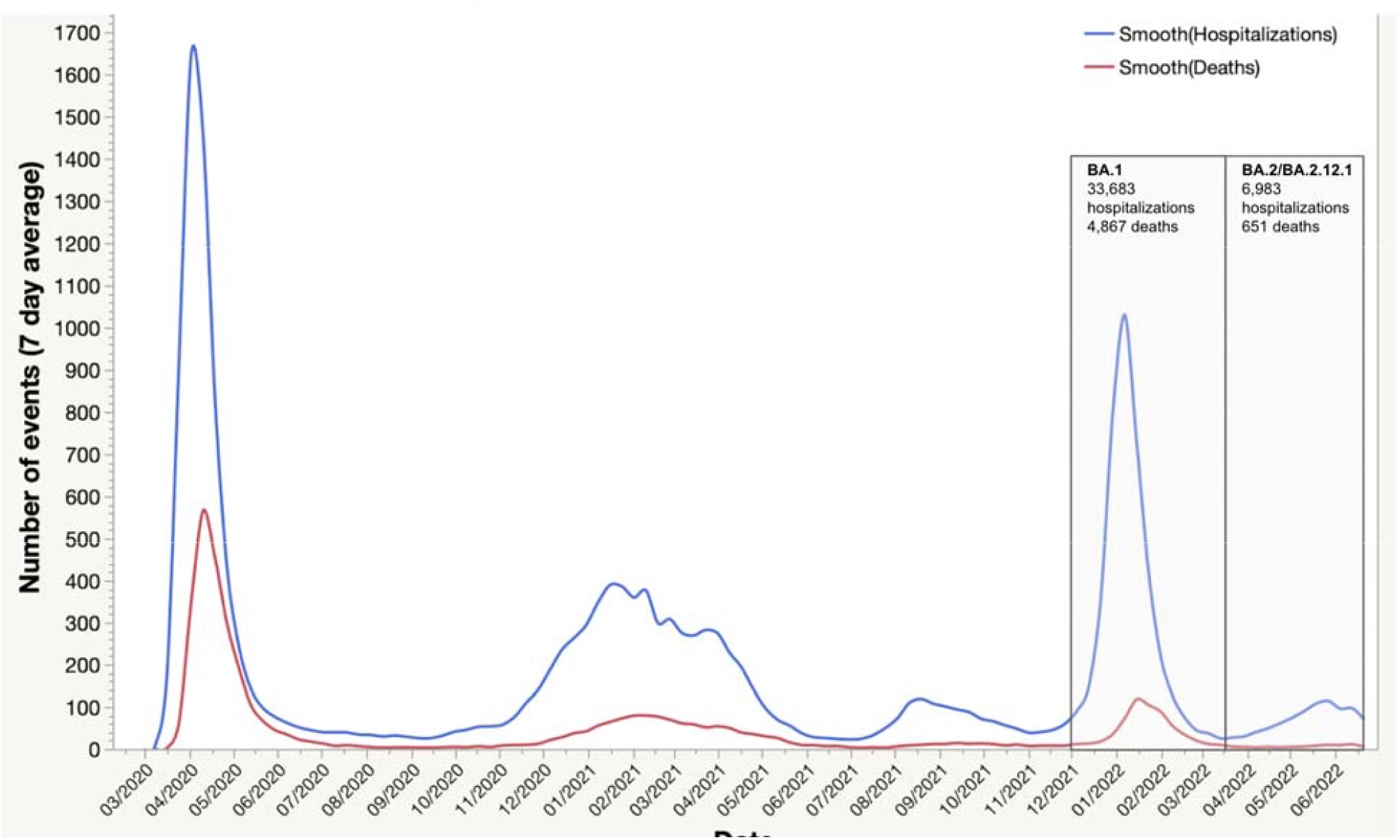
COVID-19 related hospitalizations and deaths, NYC

### Wastewater surveillance

The trend in SARS-CoV-2 wastewater concentration from the 14 NYC WRRFs is shown in Figure 4. The largest peak is for the BA.1 surge in December 2021-January 2022, followed by a more modest peak during the BA.2/BA.2.12.1 surge in March-June 2022. The BA.2/BA.2.12.1 surge was similar to the peak of the Delta surge in the fall of 2021. The ratio of SARS-CoV-2 wastewater concentration to reported cases is shown in Figure S1. The mean peak viral concentration per capita for the BA.1 surge across all 14 WWRFs was 58,294,503 N1 copies per liter per day per capita on December 26, 2021 and the peak daily case count was 60,757 on January 3, 2022 (ratio of peak wastewater concentration : peak cases=959.5). For BA.2/BA.2.12.1, these numbers were 10,743,571 N1 copies per liter per day per capita on May 3, 2022 and 5,664 cases on May 23, 2022 (ratio of peak wastewater concentration : peak cases=1,896.8), which corresponds to a two-fold higher peak concentration of SARS-CoV-2 in wastewater per reported SARS-CoV-2 case than that during the BA.1 surge.

**Figure 4.**
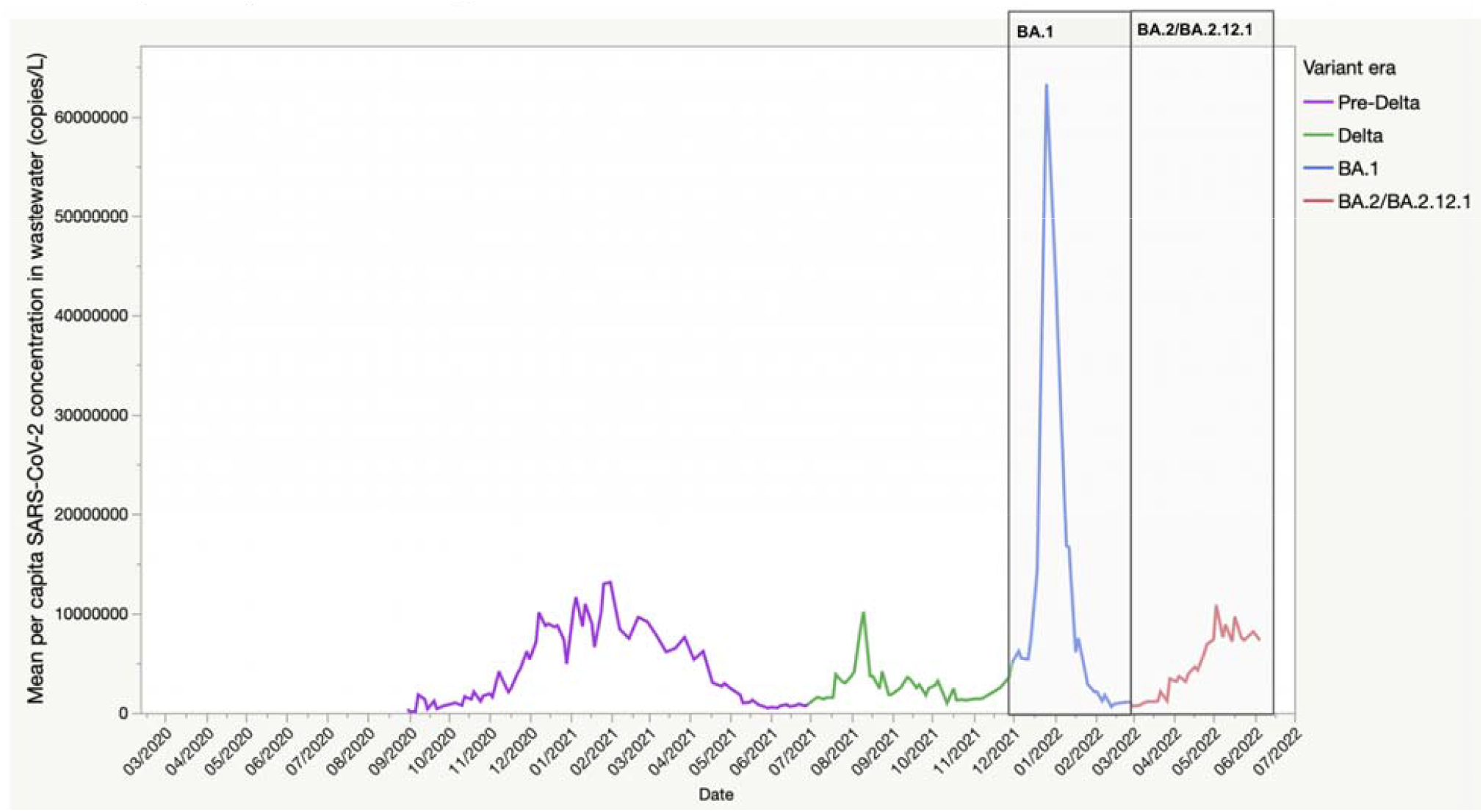
Mean per capita SARS-CoV-2 concentrations from 14 water resource recovery facilities (WRRFs) in NYC covering wastewater of an estimated 8.2 million residents.

## Discussion

Our survey found a much higher prevalence of SARS-CoV-2 infection during the BA.2/BA.2.12.1 surge in late April and early May, 2022 than was detected by surveillance. We estimate that 22.1% of adult New Yorkers, approximately 1.5 million adults, had SARS-CoV-2 infection during the two week study period, when the more transmissible BA.2.12.1 subvariant made up an estimated 20% of all cases and was increasing rapidly.^22^ The estimate of 1.5 million infections is about 29-fold higher than the 51,218 cases in the official NYC case counts^23^ and suggests a potentially vast underestimate of the BA.2./BA.2.12.1 surge’s magnitude, and a high incidence of reinfections and breakthrough infections. Importantly, while the numbers of COVID hospitalizations and deaths increased, they remained much lower than that during the recent BA.1 surge. Even though a high proportion of individuals vulnerable to a severe outcome were infected and did not use rapid antivirals, it appears that most also had protection against a severe outcome through vaccination and boosters, on top of a history of prior infection. This high degree of hybrid immunity, coupled with high vaccine- and recently acquired infection-induced immunity via BA.1, if temporary, could partly explain why NYC did not experience a major increase in hospitalizations during the BA.2.12.1 surge.

We found substantial differences in age- and sex-adjusted SARS-CoV-2 prevalence by sociodemographic factors, including race/ethnicity, which could be reflective of a number of things, alone or in combination, including greater exposure to SARS-CoV-2 (i.e., in the home, workplace or other setting(s)), and differences in individual behaviors around masking and social distancing. Household characteristics (number of household members and children) and individual behaviors may be increasingly relevant as a determinant of infection risk during surges going forward, as many pandemic restrictions had been recently dropped in NYC, leaving decisions about COVID precautions up to individual citizens.

We estimated a higher age and sex-adjusted prevalence of SARS-CoV-2 infection among those who were boosted compared with those who were fully vaccinated but not boosted and those who were unvaccinated. Since vaccines and boosters provide limited protection against *infection* with omicron, these differences are likely due to differences in SARS-CoV-2 exposures and behaviors between the two groups. These findings have important implications for observational (test negative) vaccine effectiveness (VE) studies, which are confounded by differences in exposure/behavior, testing behavior, and prior COVID between those vaccinated/boosted and unvaccinated. Given that the prevalence of prior COVID was nearly 62% among unvaccinated persons (Table 2), not taking into account the likely differential depletion of ‘susceptibles’ by vaccination status (i.e., due to SARS-CoV-2 infections) and differences in testing behaviors will bias (underestimate) VE against severe disease and death in test negative designs.^24^ Survey data such as ours can be used to correct VE estimates for these biases.

When we took into account both vaccination status and prior SARS-CoV-2 infection, we found that those with hybrid immunity had higher SARS-CoV-2 prevalence (29.2%) than those who had vaccine-induced protection only (12.9%) and a similar prevalence to those with infection-induced protection only (24.6%). This suggests that prior infection (more so than vaccination) is a strong marker for exposure risk during surges (e.g., workplace, household) and possibly reflects a lower perceived risk for infection/reinfection, severe disease/death, and onward transmission. The possible role of past and more recent SARS-CoV-2 infections in reducing adoption of personal risk mitigation measures during a surge needs to be further examined. Increasing first, second and subsequent vaccine doses among those with any history of previous infection is therefore a key strategy to help lower the population risk of severe COVID and death.

A substantial proportion (27.8%) of adults who are vulnerable to a severe SARS-CoV-2 outcome were estimated to have SARS-CoV-2, reinforcing the importance of vaccination and boosters in this group. Observational studies report that vaccine effectiveness against hospitalization with omicron (BA.1) is about 55% for two doses and 80% after a single booster, soon after dosing, and worsens substantially by 3 months.^25,26^ While recent prior infection with BA.1 does not necessarily protect against re-infection with BA.2 or BA.2.12.1^27,28^, it may confer enhanced protection against severe disease. The fraction of individuals who would likely be hospitalized with COVID-19 following recent prior omicron infection is unknown. However, in the pre-vaccine era in NYC, hospitalization would have been expected for 3.7% of re-infected individuals.^29^

Our study suggests that awareness and uptake of nirmatrelvir/ritonavir (Paxlovid™) was low among adults with SARS-CoV-2 infection in our study. Nirmatrelvir/ritonavir trials were conducted among unvaccinated individuals at high risk for hospitalization and death, and were shown to reduce the likelihood of these outcomes by ∼90%.^3^ It is unclear how much added protection nirmatrelvir/ritonavir provides over and above that provided by vaccines/boosters.

The Panoramic trial in the UK^30^ is currently assessing the efficacy of nirmatrelvir/ritonavir in fully vaccinated persons. Nonetheless, NYC recommends antivirals for individuals susceptible to severe COVID-19, regardless of vaccination status, despite the lack of evidence for an added protective effect above that provided by vaccination.^21^

Our sample size was small in analyses restricted to those with SARS-CoV-2 infection. However, stratified analyses of nirmatrelvir/ritonavir uptake by sociodemographic factors and biologic vulnerability suggested that there may be important inequities in antiviral access across a number of social determinants of health. A recent national CDC study had similar findings.^31^ While caution is warranted in the interpretation of our estimates of these inequities, as the confidence limits were wide, these are potentially important findings that warrant further investigation and monitoring, with policy and programmatic course corrections as needed. Inequitable uptake of antivirals among vulnerable individuals with COVID-19 will further exacerbate inequities in the burden of SARS-CoV-2 which has had disproportionate effects on racial/ethnic minorities and other groups.^32–34^ It is essential to give attention to both need and equity in the design and implementation of large scale public health initiatives, and to avoid designs were such initiatives may create new inequities or exacerbate existing inequities.

Despite our estimate of a surge in infections among adults during the two week study period, the signal of SARS-CoV-2 concentration per capita in NYC wastewater surveillance data, which covers an estimated 8.2 million New Yorkers, was modest in comparison to that during the BA.1. surge. When we accounted for the number of reported cases, the mean SARS-CoV-concentration per reported case in wastewater was twice as high during the BA.2.12.1 surge vs. BA.1. This could reflect an increase in the mean peak viral load per case between the two subvariant surges (e.g., a result of the properties of the pathogen and/or host susceptibility), and/or a decline in the completeness of case reporting coupled with an increased reliance on at-home rapid testing. The latter seems a more plausible explanation, given the similarities of BA.1 and BA.2/BA.2.12.1. Rather, because of recent prior BA.1 infection among a wide swath of the NYC adult population, it may be that those re-infected with BA.2/BA.2.12.1 after a recent BA.1 infection had a *lower* mean peak viral load due to greater acquired T-cell mediated immunity from prior BA.1 infection.^35–37^ This could effectively reduce the magnitude of peak SARS-CoV-2 concentration in wastewater, even for roughly the same number of individuals in the population with active SARS-CoV-2 infection. If this is true, it would suggest that wastewater surveillance may be less useful for surge detection soon after a recent widespread surge like BA.1, and requires further investigation. It would also suggest that the public health benefits of vaccination and hybrid immunity may be even greater than currently appreciated.

Indeed, a very large pre-Omicron study from Qatar compared cycle threshold (Ct) values of those with active primary, reinfection, or breakthrough infection, adjusting for sex, age, reason for testing and calendar week of testing. Compared with unvaccinated people with primary infection, Ct values were 4.0 cycles higher in unvaccinated people who had reinfection, followed by vaccinated people with a breakthrough infection (1.3 cycles higher for BNT162b2 breakthrough infections and 3.2 cycles higher for those with mRNA-1273 breakthrough infections).^38^ A more recent study of those with BA.1 or BA.2 infection found that while those with BA.2 had adjusted cycle thresholds that were 3.5 cycles lower compared with BA.1, having a prior infection <90 days before a reinfection was associated with adjusted cycle thresholds that were 4.23 cycles higher.^39^ These studies suggest that peak viral loads and/or length of shedding is lowest in those with a reinfection. There are no equivalent studies for breakthrough infections among those who had both a prior infection and were vaccinated (hybrid immunity), but it is possible that cycle thresholds could be higher.

We found a disconnect between the number of tests and positive tests with a healthcare provider in our survey and those reported in official SARS-CoV-2 surveillance data. About 43% of adults in our survey reported having received a SARS-CoV-2 test with a health care or testing provider in the prior 2 weeks, which would correspond to about 2.9 million adults tested during April 23-May 8, 2022. However, according to NYC surveillance data, only ∼670K tests were performed during the study period^23^, suggesting that testing itself (and possibly rapid testing specifically) may be severely underreported by laboratories and providers (by a factor of 4), overestimated by our survey, or a combination of both. Passive surveillance relies on institutions to voluntarily report data (often in batches using electronic systems). However, data quality, timeliness and completeness cannot be guaranteed and can be variable. If tests, including positive tests, are underreported, this could be part of the reason for the larger than expected discrepancy between case counts and our estimate of SARS-CoV-2 infections. Unfortunately, our survey (Appendix 2) did not distinguish between PCR and rapid tests with provider-based testing. However, in a recent EHR based analysis of data from a large NY area urgent care provider, our team found that rapid antigen tests were more common than PCR tests, including among positives^40^, which conflicts with city-wide data on testing. To our knowledge, the completeness, representativeness, timeliness and acceptability of passive SARS-CoV-2 reporting of such a high volume of cases and test results by providers and laboratories, including during surges, has not been systematically assessed in NYC or elsewhere around the U.S., making it important to investigate this discrepancy. A recent analysis of Omicron infections over a five day period (December 13-17, 2021) in England from the REACT-1 study estimated that the community SARS-CoV-2 prevalence (based on PCR swab positivity in study participants) was about 600,000, and this compared with 206,295 confirmed and probable Omicron cases diagnosed and reported through routine case surveillance from December 1-December 21, suggesting substantial under ascertainment of cases in a setting where there are functional national systems for capturing the results of provider-based testing.^41^ In general, passive surveillance of infectious diseases in the U.S. that relies on reporting by health care providers has low completeness, especially when there is an administrative burden to complete forms or enter data.^42^ However, laboratory reporting (i.e., of tests and related results) that leverages laboratory information systems can be higher. Completeness of both types of reporting is challenging to assess and improve. However, during public health emergencies, completeness can be enhanced via increased awareness and reminders to providers and laboratories around reporting obligations. Large surges in cases could create added administrative burden and demand on providers and laboratories, potentially reducing the proportion of tests and cases reported or reducing data quality/completeness. This may be particularly true for point of care tests during intense surges. Of note, effective April 4, 2022, HHS, CDC and New York State announced changes to laboratory reporting requirements that no longer required reporting of negative or inconclusive rapid antigen tests.^43^

Our survey estimates of SARS-CoV-2 prevalence and provider testing could be inflated if those who both tested for SARS-CoV-2 infection and tested positive were more likely to participate in the survey than those who did not. While potential survey participants were not aware of the survey content before deciding to participate, it may be that those who were positive were more likely to complete the survey. We could neither rule out nor correct for this non-response bias, however, characteristics of survey respondents did not differ substantially from that of the adult NYC population (Appendix 1). It is also possible that participants inadvertently recalled and reported positive tests that were beyond the 14 day study period (recall bias). Lastly, some people test multiple times with providers after their initial positive test^40^, and subsequently, many can expect positive PCR and antigen test results for 10 or more days.^44,45^ This could have caused some people who were diagnosed prior to the study period to have positive tests during the study period which could have inflated our prevalence estimates relative to official case counts. But from an epidemiologic standpoint, these individuals should be reflected in prevalence estimates as they are actively infected.

Our study had other limitations, including a limited sample size, especially in subgroups of those with evidence of COVID-19. For those with prior COVID, we did not capture information on timing of prior infections, which may overestimate the degree of current hybrid immunity, though a substantial proportion of NYC adults were infected during the recent BA.1 surge.^9,23^ Our case definition would likely capture some of the estimated 20-30% of individuals whose SARS-CoV-2 infection may remain asymptomatic throughout their infection,^46,47^ as well as those who were symptomatic but were not aware of a close contact. Finally, our survey did not include children or those whose primary language was not English or Spanish.

Strengths of our study include the representative and probability-based design of the survey, the ability for the survey to reflect outcomes among those who do not access the healthcare system, and triangulation with other important data sources such as routine passive surveillance for cases, hospitalizations, and deaths, as well as wastewater surveillance. Other strengths include the study’s timing at the start of the BA.2/BA.2.12.1 surge, and measurement of several important factors that are not currently available through routine surveillance, including outcomes among those who do not test at all or test exclusively at home during a surge, prevalence among individuals vulnerable to COVID-19, hybrid immunity, and awareness/uptake of nirmatrelvir/ritonavir.

## Conclusions

Our study’s findings suggest that the magnitude of NYC’s BA.2/BA.2.12.1 surge was underestimated by official case counts and wastewater surveillance, due to a combination of exclusive at-home testing, not testing at all, incomplete provider/laboratory reporting, and recently acquired omicron infection (BA.1). This is likely also true for other U.S. jurisdictions with similar approaches, and by extension, the national SARS-CoV-2 surveillance system. Even though many individuals vulnerable to a severe outcome were infected and did not use rapid antivirals, it appears that most had a high degree of protection against a severe outcome through vaccination and boosters, overlaid by a recent history of prior BA.1 infection, limiting the surge’s impact on hospitalizations and deaths. However, given the uncertainty at the outset of how surges will impact severe outcomes among those who remain susceptible, a shift in approach to public health surveillance for SARS-CoV-2 is needed for timely surge detection and quantification in the U.S. Our findings demonstrate the utility of population-representative surveys as an important surveillance tool to go alongside, and triangulate, with passive case reporting, genomic surveillance, and wastewater surveillance at this uncertain and evolving stage of the U.S. pandemic.

## Data Availability

All data produced in the present study are available upon reasonable request to the authors.

## Data Availability

All data produced in the present study are available upon reasonable request to the authors.

## Acknowledgements

The authors wish to acknowledge the survey participants and Consensus Strategies for completing survey sampling and data collection.

---

**Figure S1.**
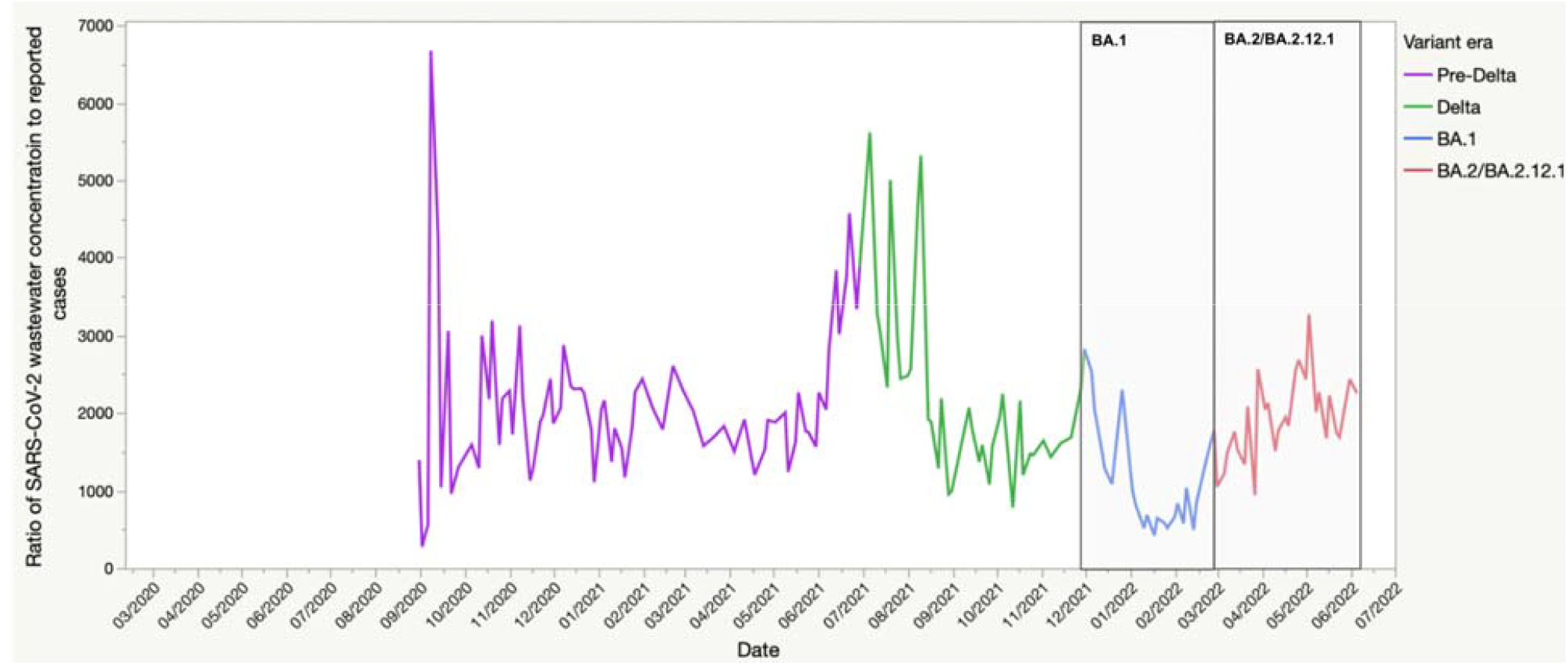
Ratio of SARS-CoV-2 concentration in NYC wastewater to reported SARS-CoV-2 cases

### Appendix 1 (Survey design)

#### Sampling Frame

A sampling frame of 5,580,901 residents of New York City was used of which 2,185,433 have mobile numbers with an additional 1,539,793 landlines. Two stratified proportionate randomized population-based samples were drawn for this study, n=45,000 mobile numbers and n=60,000 landlines. A total sample of n=1,030 was utilized with a +/- 3 percent margin of error.

#### Multi-mode data collection design

Short message service (SMS) aka text messages were sent using SMS platform. The respondents were sent a personalized first name text message which included a link to the survey and an opt-out option. The respondents had the option to reply to the SMS text with any queries and the survey was available in both English and Spanish. Data was verified by IP address and scrubbed against the original survey sample.

Interactive voice response (IVR) aka robo-poll messages were sent to landlines. The respondents were able to answer the survey questions using the touch tone keypad on their phones in either English or Spanish.

Data were collected on May 7 and 8, 2022.

#### Response rates

In the table below, we present the response rates for our survey by modality. For context, we also present response rates and participation rates for the same modalities for New York City’s 2020 Community Health Survey (CHS) and the NYC Health Opinion Poll (HOP) in Q1 (HOP1) and Q2 (HOP2) in 2019. While our response and participation rates are low, population-based surveys, including those in NYC, have seen increasingly low response rates (Table). The higher response rates for the CHS survey relative to our BA.2 survey could be due to publicity and outreach efforts that are conducted by the NYC Department of Health in order to increase participation. Lower response rates in our survey could be due to the fact that it was conducted during a COVID surge when people may have been less likely to be available to participate in surveys. Importantly, population-based surveys in the US have been seeing increasingly low response rates but as a <u>recent paper by the Pew Research Center notes</u>, they can still provide accurate data with low response rates. To assess the impact of non-response on study validity, studies that have examined non-response in surveys suggest that response rates are a poor indicator of non-response bias and data quality.

|  | NYC BA.2 Survey (current survey) |  |  | 2020 Community Health Survey (CHS) |  |  | NYC HOP-1 (2019) | NYC HOP-2 (2019) |
| --- | --- | --- | --- | --- | --- | --- | --- | --- |
| Modality | Landline | Cell | Combined | Landline | Cell | Combined | Online opt-in panel | Online opt-in panel |
| Response rate (%) | 1.3 | 0.6 | 1.2 | 6 | 7.8 | 7.4 | -- | -- |
| Participation rate (%) | 21.6 | 37.6 | 24.2 | 68.4 | 76 | 74.4 | 1.9 | 2.3 |

#### Comparison of characteristics between complete respondents and adults in New York City based on the American Community Survey, 2020

To assess whether there may have been systematic response bias, we compared the unweighted distributions of those completing the survey to that of adult NYC residents from the 2020 American Community Survey (ACS). We did not find large discrepancies between the survey sample and NYC population except that our survey respondents skewed slightly older, which is likely due to older adults being available to respond to the survey. Importantly, in our study, potential respondents were not informed in advance that the survey would be about COVID, but rather told that the survey was focused on public policy.

| ***Race/ethnicity*** | ***Survey Completers*** | ***ACS NYC 2020*** |
| --- | --- | --- |
| *White NH* | *39.8* | *31.9* |
| *Black NH* | *20.7* | *21.4* |
| *Hispanic* | *28.5* | *28.9* |
| *AAPI* | *6.4* | *14.2* |
| *Other/more than one race* | *4.7* | *3.5* |

| ***Age*** | ***Survey Completers*** | ***ACS NYC 2020*** |
| --- | --- | --- |
| *18-24* | *5.1* | *8.3* |
| *25-34* | *8.7* | *17.8* |
| *35-44* | *6.8* | *13.6* |
| *45-54* | *8.8* | *12.5* |
| *55-64* | *17.0* | *12.1* |
| 65+ | 53.6 | 15.0 |

| **Gender** | **Survey Completers** | **ACS NYC 2020** |
| --- | --- | --- |
| *Male* | 41.5 | 47.7 |
| *Female* | 55.1 | 52.3 |
| *Non-binary* | 3.5 | -- |

| **Education** | **Survey Completers** | **ACS NYC 2020** |
| --- | --- | --- |
| *Less than HS* | 12.2 | 16.7 |
| *HS graduate* | 21.4 | 23.7 |
| *Some college* | 25.2 | 22.4 |
| *College grad* | 41.2 | 37.1 |

#### Weighting

The survey was weighted using an iterative weighting method (raking) to marginal proportions of race, ethnicity, age, gender and education by borough. The sum of the weights equals the sample population (n=1030). The demographic weights were developed based on the American Community Survey ANNUAL ESTIMATES OF THE RESIDENT POPULATION: APRIL 1, 2010 TO JULY 1, 2019 - FOR FULL ESTIMATES DETAIL, VISIT: https://www.census.gov/programs-surveys/popest.html

The weights were based on respondent self-identified sex, educational attainment, age, ethnicity/race and region. The inference population is 6,740,580 million adult NYC residents.

### Appendix 2 (Survey questionnaire)

#### Survey on recent COVID exposure, COVID infection, and testing behaviors in New York City

Hello, this is XYZ with a brief public policy survey. At no time will we try to sell you anything. We are just interested in your opinions, and you can drop out at any time.

To begin, what language would you like to take this survey in?

a. English
b. Español

##### The following questions will ask about COVID exposure in the past 2 weeks

1. In the past 2 weeks, have you experienced any COVID-like symptoms (e.g., 100 degrees fever or higher, chills, cough, sore throat, fatigue, headache, shortness of breath, congestion or runny nose, muscle aches, loss of smell or taste, nausea, or diarrhea)?
  a. Yes
  b. No
  c. Don’t know/not sure
2. In the past 2 weeks, has anyone in your household (not including yourself) experienced COVID-like symptoms or tested positive for COVID-19?
  a. Yes
  b. No
  c. Don’t know/not sure
3. In the past 2 weeks, were you exposed to any other person outside of your household who had COVID-like symptoms or tested positive for COVID-19?
  a. Yes
  b. No
  c. Don’t know/not sure

##### The following questions will ask about COVID testing and treatment in the past 2 weeks

4 In the past 2 weeks, have you taken an at-home rapid test for COVID-19? (a rapid at-home test allows you to collect your own sample and get results within minutes at home)
  a. Yes, Tested Positive
  b. Yes, Tested Negative
  c. No, I have not tested
5 In the past 2 weeks, have you taken a rapid antigen or PCR test for COVID-19 from a healthcare or testing provider?
  a. Yes, Tested Positive (go to 6)
  b. Yes, Tested Negative (go to 6)
  c. No, I have not tested (go to 9)
6 In the past 2 weeks, which of the following locations did you get tested for COVID-19?
  a. Hospital or physician’s office
  b. Urgent care clinic
  c. Pharmacy
  d. Mobile testing site
  e. Employer
  f. Other
7 In the past 2 weeks, how difficult was it for you to get yourself a viral COVID-19 test at a healthcare or testing provider (PCR or rapid) if you attempted to get one?
  a. Did not attempt to get a test
  b. Not Difficult
  c. Somewhat Difficult
  d. Very Difficult
8 In the past 2 weeks, did you try to get paxlovid, an antiviral medication for COVID-19?
  a. No, I don’t know about paxlovid
  b. No, I did not try to get a paxlovid prescription
  c. Yes, I received a paxlovid prescription
  d. Yes, I tried to get paxlovid, but was unable to get it
9 Have you ever self-administered an at-home rapid test for COVID-19 for yourself or for someone in your household?
  a. Yes
  b. No
  c. Don’t know/not sure
10 If you had easy access to free at-home rapid tests, would you prefer to test for COVID-19 at using at-home rapid tests or at a health care/test center?
  a. Take at-home rapid test
  b. Health Care/Test Center
  c. Don’t know/not sure

##### Do you agree or disagree with the following statements about COVID-19 at-home testing?

11 At-home rapid tests are easy to use:
  a. Strongly agree
  b. Agree
  c. Neither agree nor disagree
  d. Disagree
  e. Strongly disagree
12 I can easily get an at-home rapid test if I or someone in my household needs one:
  a. Strongly agree
  b. Agree
  c. Neither agree nor disagree
  d. Disagree
  e. Strongly disagree
13 Since March 2020, have you ever had COVID-19 infection and tested positive, either at home or with a provider?
  a. Yes, once
  b. Yes, more than once
  c. No, but I am pretty sure that I had COVID
  d. No, I never tested positive, and I don’t think I have ever had COVID
  e. Don’t know/not sure
14 Were you aware that New York City just moved from the low to the medium COVID-19 risk level, indicating higher levels of community transmission?
  a. Yes
  b. No
  c. Don’t know/not sure

##### Respondent Characteristics

15 Do you currently have any kind of health care coverage, including health insurance, prepaid plans such as HMOs, or government plans such as Medicaid or Medicare, or Indian Health Service?
  a. Yes
  b. No
  c. Don’t know/not sure
16 Do you have any of the following conditions that could increase the severity of COVID-19: cancer, diabetes, obesity, COPD or lung disease, liver disease, heart disease, high blood pressure, a recent organ transplant, or an immunodeficiency)?
  a. Yes
  b. No
  c. Don’t know/not sure
17 17. Have you been fully vaccinated against COVID-19? [Either 2 doses of mRNA vaccine series (Moderna or Pfizer) or a single dose of Johnson and Johnson COVID-19 vaccine]
  a. Yes (go to 18)
  b. No (go to 19)
  c. Don’t know/not sure (go to 19)
18 If you have been fully vaccinated, have you also received a coronavirus booster?
  a. Yes, more than 5 months ago
  b. Yes, within the past 5 months
  c. No
19 If not fully vaccinated OR not boosted: Do you plan to get a vaccine dose or booster in the next two weeks?
  a. Yes
  b. No
  c. Don’t know/not sure
20 Which zip code do you reside in?
21 What is your age?
  a. 18-24
  b. 25-34
  c. 35-44
  d. 45-54
  e. 55-64
  f. 65-74
  g. 75 +
22 How do you currently identify your gender? Do you identify as …
  a. Male
  b. Female
  c. Gender non-binary
23 Are you Latino/a, or of Hispanic or Spanish origin?
  a. Yes
  b. No
24 Which one of the following would you use to describe yourself ?
  a. White
  b. Black or Black American
  c. Asian, Native Hawaiian or Other Pacific Islander
  d. More than one race
25 Were you born in the USA [Puerto Rico and other territories are considered outside the USA]?
  a. Yes
  b. No
  c. Don’t know
26 What is the highest grade or year of school you completed?
  a. Less than high school
  b. Grade 12 or GED (High school graduate)
  c. College 1 year to 3 years (Some college or technical school, associate degree)
  d. College 4 years or more (College graduate)
27 How many members of your household, including yourself, are 18 years of age or older?
  a. 1
  b. 2
  c. 3
  d. 4
  e. 5
  f. 6
  g. 7
  h. 8
  i. 9 Or more
28 How many children 17 years old or younger usually live or stay with you?
  a. 0
  b. 1
  c. 2
  d. 3
  e. 4
  f. 5
  g. 6
  h. 7
  i. 8
  j. 9 Or more
29 Are you currently employed for wages or salary?
  a. Yes
  b. No
  c. Don’t know/not sure
30 What is your household’s annual income?
  a. $25,000 or less
  b. Between $25,001 - $65,000
  c. Between $65,000 - $150,000
  d. Above 150,000
  e. Refuse

